# The Role of Artificial Intelligence in Modern Medical Education and Practice: A Systematic Literature Review

**DOI:** 10.1101/2024.07.25.24311022

**Authors:** Shiva Rasouli, Duha Alkurdi, Bochen Jia

## Abstract

The integration of Artificial Intelligence (AI) into medical education has emerged as a transformative element in the modern healthcare educational system. With the exponential growth of medical knowledge and the increasing complexity of healthcare systems, AI offers innovative solutions to enhance learning outcomes, facilitate personalized education pathways, and improve clinical decision-making skills among medical professionals. This literature review explores the transformative role of AI in the training of healthcare providers, focusing on advancements in medical education, medical diagnostics, and emergency care training. Additionally, it addresses the readiness of healthcare professionals to employ AI technologies, analyzing their current knowledge, attitudes, and the training provided. By synthesizing findings from multiple studies, we aim to highlight AI’s potential to enhance medical education, address challenges, and propose future directions for integrating AI into healthcare training.

## 1 Introduction

### 1.1 Background

The field of medicine is inherently complex and continuously evolving, driven by a constantly expanding knowledge base. Advances in medical research, technological innovations, and shifts in healthcare practices contribute to this growth. As new diseases are identified and innovative treatments are developed, medical education becomes increasingly intricate. It is crucial for medical professionals to not only acquire extensive knowledge during their education but also to keep up with the latest developments, techniques, and treatments throughout their careers. Ongoing education is vital for delivering effective patient care as well as ensuring that practitioners remain competent and relevant in this fast-paced, ever-changing field.

Traditional medical education, which has been grounded in classroom learning, hands-on training, and mentorship, focuses on six fundamental areas: patient care, medical knowledge, communication skills, continuous learning, professionalism, and systemic practice [1, 2, 3]. This education system prioritizes deep knowledge acquisition, heavily relying on memorization to prepare students for patient care [4]. Traditional methods, such as classroom instruction, often fall short of capturing the complex, unpredictable nature of real-life medical scenarios. Hands-on training, though valuable, is frequently constrained by resource availability, from patient access to practical opportunities such as in surgery [5]. Additionally, the variability in mentorship quality, dependent on a mentor’s expertise and time, can affect the consistency and depth of a medical student’s learning experience. As we advance, integrating Artificial Intelligence (AI) and other technological innovations into medical training could revolutionize educational methods, making them more adaptive, efficient, and reflective of contemporary medical practices.

AI is recognized for its ability to replicate tasks that usually require human intelligence, such as analyzing data and making decisions [6]. One of its key advantages is efficiency; AI can process and analyze extensive datasets much faster than humans. Additionally, AI operates continuously, with no human limitations like fatigue, ensuring high accuracy and speed. This is particularly beneficial in healthcare education, where AI can facilitate personalized learning, simulate clinical scenarios for training, and update educational content based on the latest research and clinical practices. These capabilities make AI a powerful tool in enhancing the training of healthcare professionals, leading to improved patient care and innovative medical practices.

AI-based methods are revolutionizing the way medical professionals learn, practice, and perfect their skills [7, 8, 9]. AI can provide a scalable and consistent educational experience, mitigating the limitations of traditional mentorship and resource-dependent training [10]. Virtual simulations and AI-driven virtual patients, for instance, offer medical students and professionals the opportunity to practice and improve their skills in a controlled environment [11, 12, 13, 14]. A study in Johns Hopkins Bayview Medical Center in Baltimore, Maryland, found that just 9 hours of deliberate practice with real-world virtual patient simulations significantly increased the diagnostic skills of internal medicine interns in evaluating dizziness [15]. These simulations can be designed to mimic a wide range of medical conditions and scenarios, providing a diverse and comprehensive training experience that might be difficult to achieve in the real world. Moreover, AI algorithms that assist in reading radiological images, are not only used in clinical settings but also as a teaching tool, enabling students to compare their analyses with AI-generated interpretations [16].

The integration of AI into medical training represents a transformative shift not only in educational methodologies but also in the preparation of medical professionals for a technologically advanced future [17, 18, 19, 20]. In contemporary clinical practice, AI and other digital tools are becoming foundational components, reshaping patient care, diagnostics, and treatment planning. This evolution necessitates a parallel transformation in medical education, where traditional training methods are increasingly supplemented with AI-driven technologies. By integrating AI into the curriculum, educational institutions aim to equip future medical professionals with the necessary skills to utilize these advanced technologies effectively. Familiarity with AI-driven tools, sophisticated data analysis techniques, and digital health applications is crucial, as these competencies will become central to the efficacy of healthcare delivery.

This literature review critically assesses the integration of AI in medical education and practice. It focuses on providing a comprehensive analysis of how AI tools are currently used and their impact on learning and patient care. Moreover, it explores what strategies can be adopted to foster the readiness of healthcare providers for utilizing AI. By gathering and evaluating evidence from various studies, this review will help educators, healthcare professionals, and policymakers make informed decisions about effectively incorporating AI technologies into medical education and practice.

### 1.2 Research Purpose

Given the complexity and dynamic nature of the medical field, where continuous learning and adaptation are crucial, the role of AI in healthcare education is increasingly important.

This literature review paper aims to shed light on the role of AI in training healthcare providers. Specifically, we seek to address these research questions:

1. How is AI utilized as an innovative educational and practice solution for healthcare professionals?
2. What strategies can be employed to foster readiness to use AI among healthcare professionals?

The first question aims to identify and explore the various ways AI is being used in medical education. The significance of this inquiry lies in its potential to reveal innovative methods for training healthcare professionals. By understanding how AI is currently utilized, we can identify best practices, gaps, and opportunities for further innovation. This can lead to more effective, efficient, and engaging training methods, ultimately enhancing the skill set of healthcare providers. The second question addresses the challenge of integrating new technologies into the healthcare sector. The significance of this question lies in the understanding that the successful implementation of AI in medical training requires not only technological solutions but also the readiness and engagement of healthcare professionals. By identifying strategies to promote readiness, this research can contribute to smoother and more effective integration of AI in medical education. This is key to ensuring that healthcare professionals are equipped to work alongside emerging technologies and leverage them for improved patient care.

The specific objective of this systematic review is strategically designed to comprehensively assess the impact of AI in medical education.

## 2 Methods

### 2.1 Design and Search

In conducting the systematic literature review, a comprehensive search was carried out across three major databases: Scopus, Google Scholar, and PubMed. Utilizing three distinct searching syntaxes, ***(“Nurse” OR “Patient” AND “Training” AND “AI”), (“Artificial Intelligence” AND “Health Care” AND “Training” AND “Patient”)*** and ***(“Artificial Intelligence” AND “Health Care” AND “Training”)*** a pool of 295 papers was initially identified, followed by a two steps screening. The first step involved screening abstracts to quickly identify and exclude studies that clearly did not meet the inclusion criteria. The second step involved a methods review of the studies that were included after the first screening step to confirm their relevance and quality in addressing the research questions and meeting all inclusion criteria. These steps resulted in 32 papers being considered for the full review.

### 2.2 Inclusion & Exclsuion Criteria

Studies that discuss the use of AI in medical education or training, including its impact on learning processes and effectiveness were included in this literature review. Additionally, research that investigates the readiness of healthcare practitioners to adopt AI, examining their attitudes and knowledge, was included. To ensure a comprehensive understanding, both qualitative and quantitative studies have been included to capture diverse methodologies and insights. These criteria help to concentrate on how AI is integrated into medical education and the acceptance of this technology in the healthcare sector.

Specific exclusion criteria were established to maintain the focus and relevance of the literature review. First, papers not written in English were excluded to ensure that all data was accessible and clear. Second, studies that examine the use of AI in domains other than healthcare were omitted, as the research specifically targets AI’s implications for medical professionals. These criteria helped streamline our review by filtering out studies that did not align with our research objectives.

## 3 Results & Discussion

Following the initial round of paper extraction, a total of 295 papers were identified, those papers met the searching syntaxes across the three platforms. Subsequently, 227 papers were excluded during the first screening process as they did not meet the inclusion criteria. During the methods review process, the count of papers was narrowed down to 32. A summary of these findings is presented in the PRISMA chart depicted in Figure 1.

**Figure 1:**
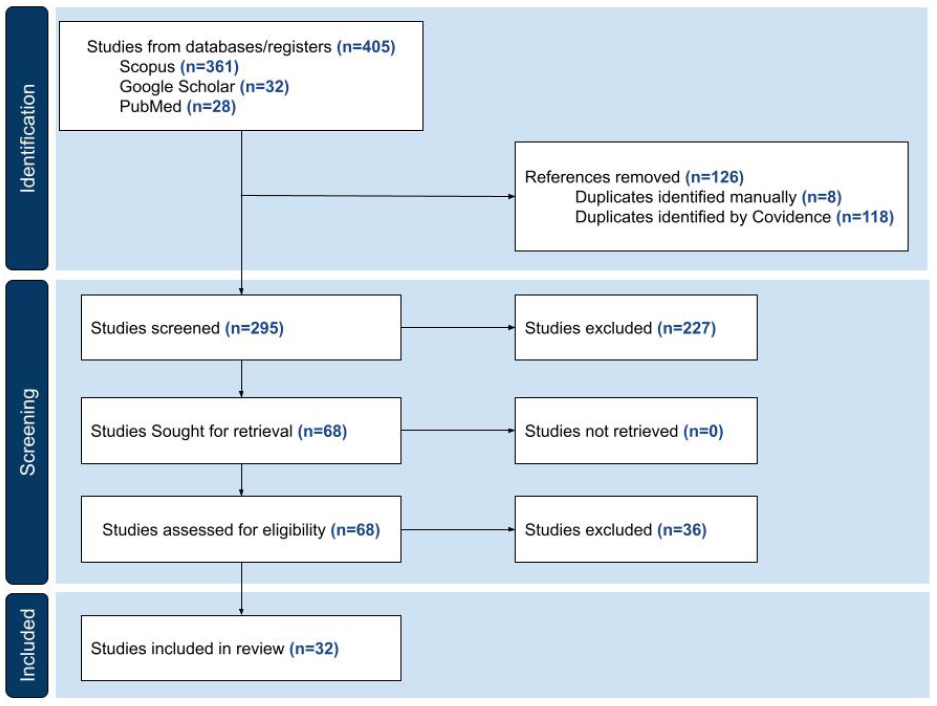
PRISMA chart

This literature review paper aims to shed light on the role of AI in training healthcare providers. Specifically, seeks to address two research questions. Table 1 presents a summary of the 32 papers included in this literature review, outlining their main objectives, implications, study results, and limitations.

**Table 1:**
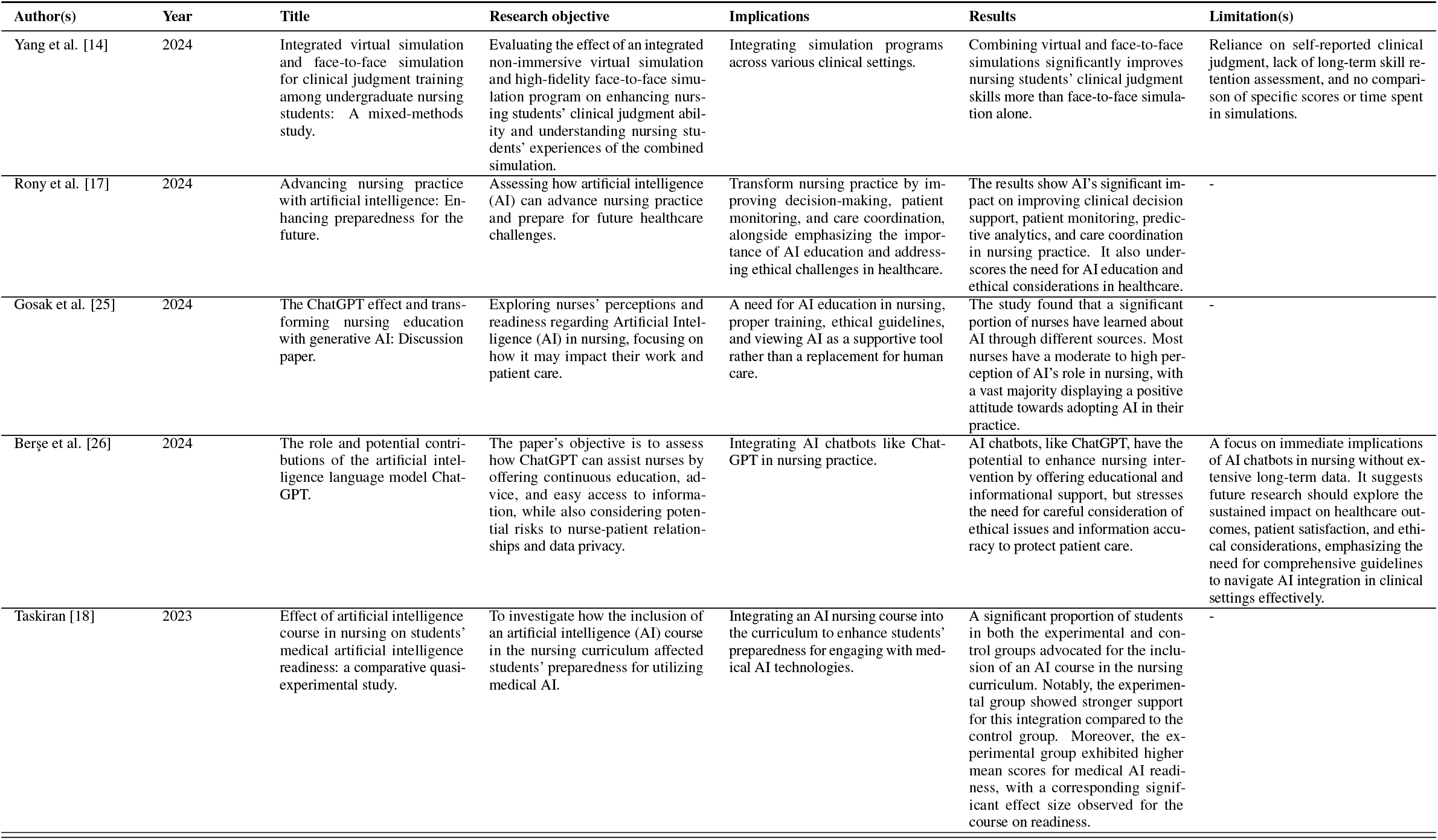
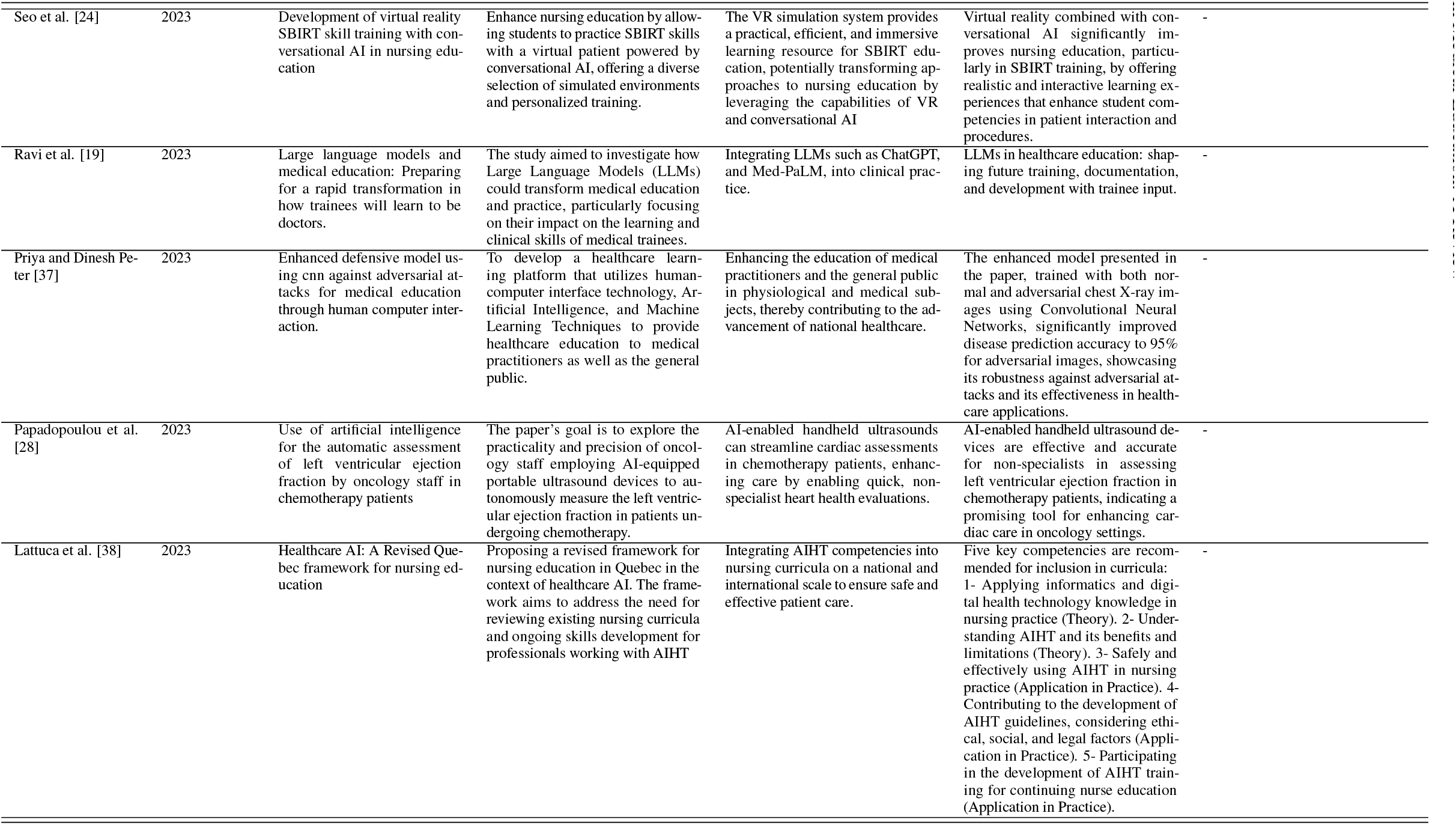
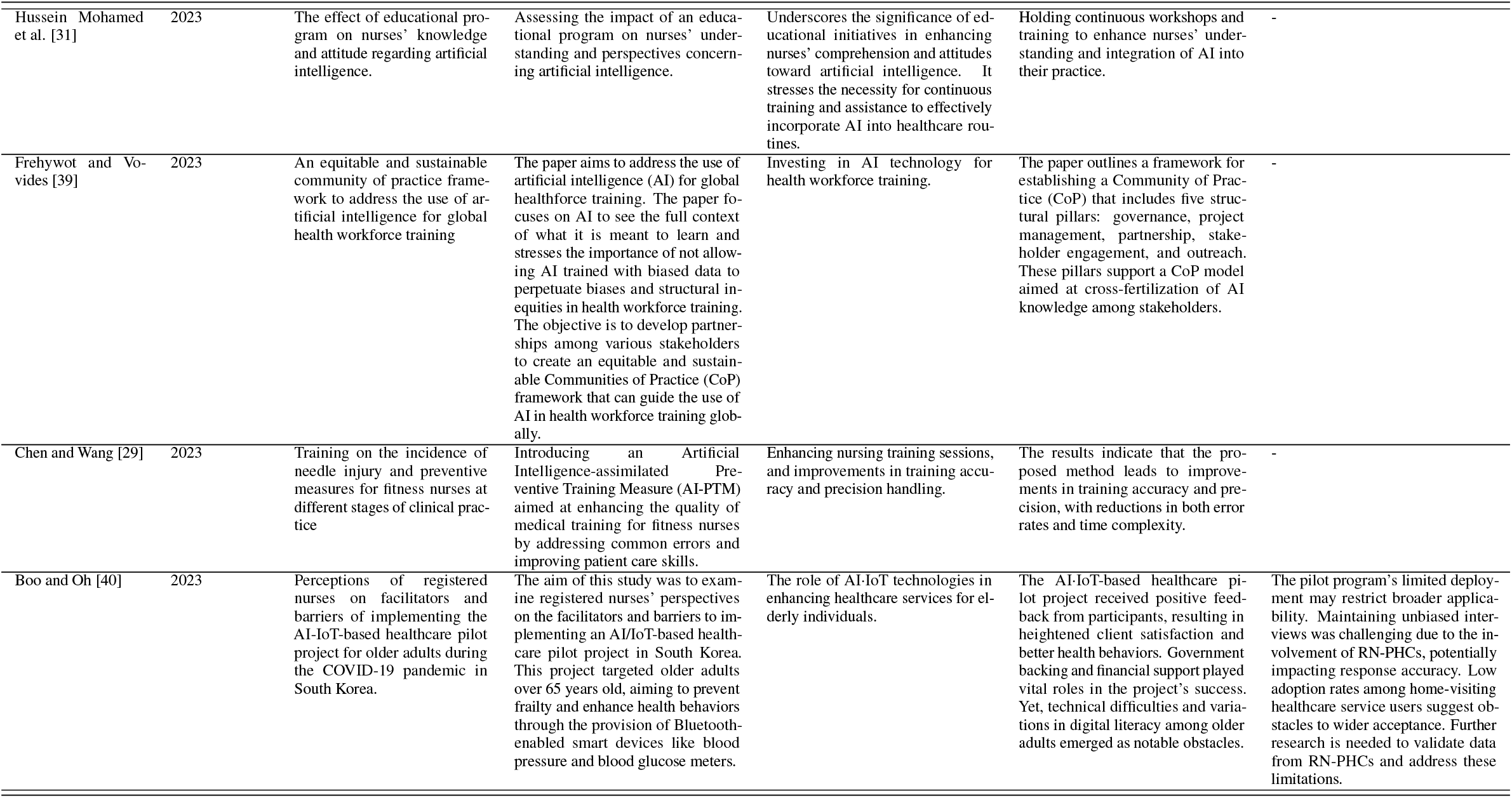
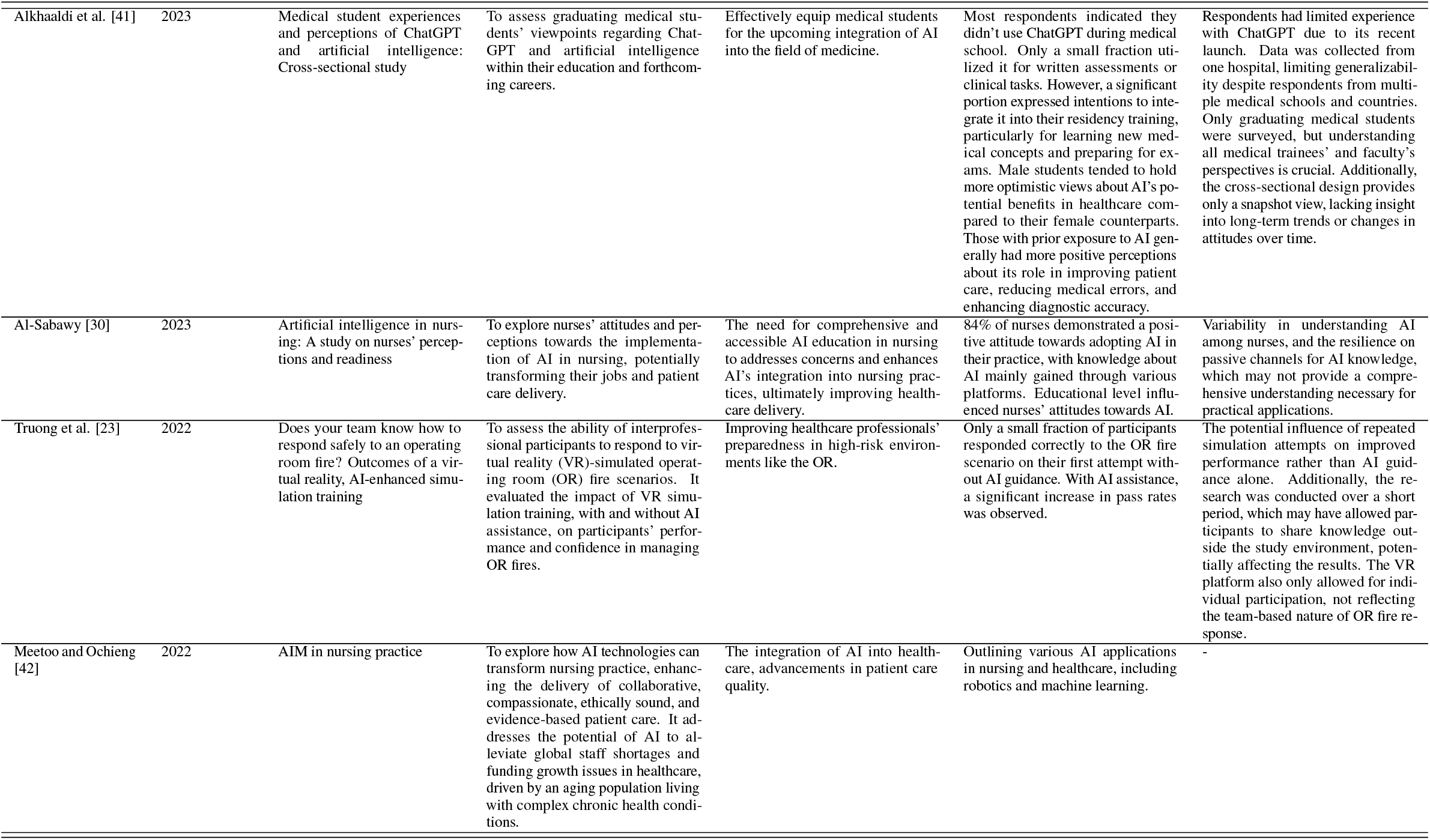
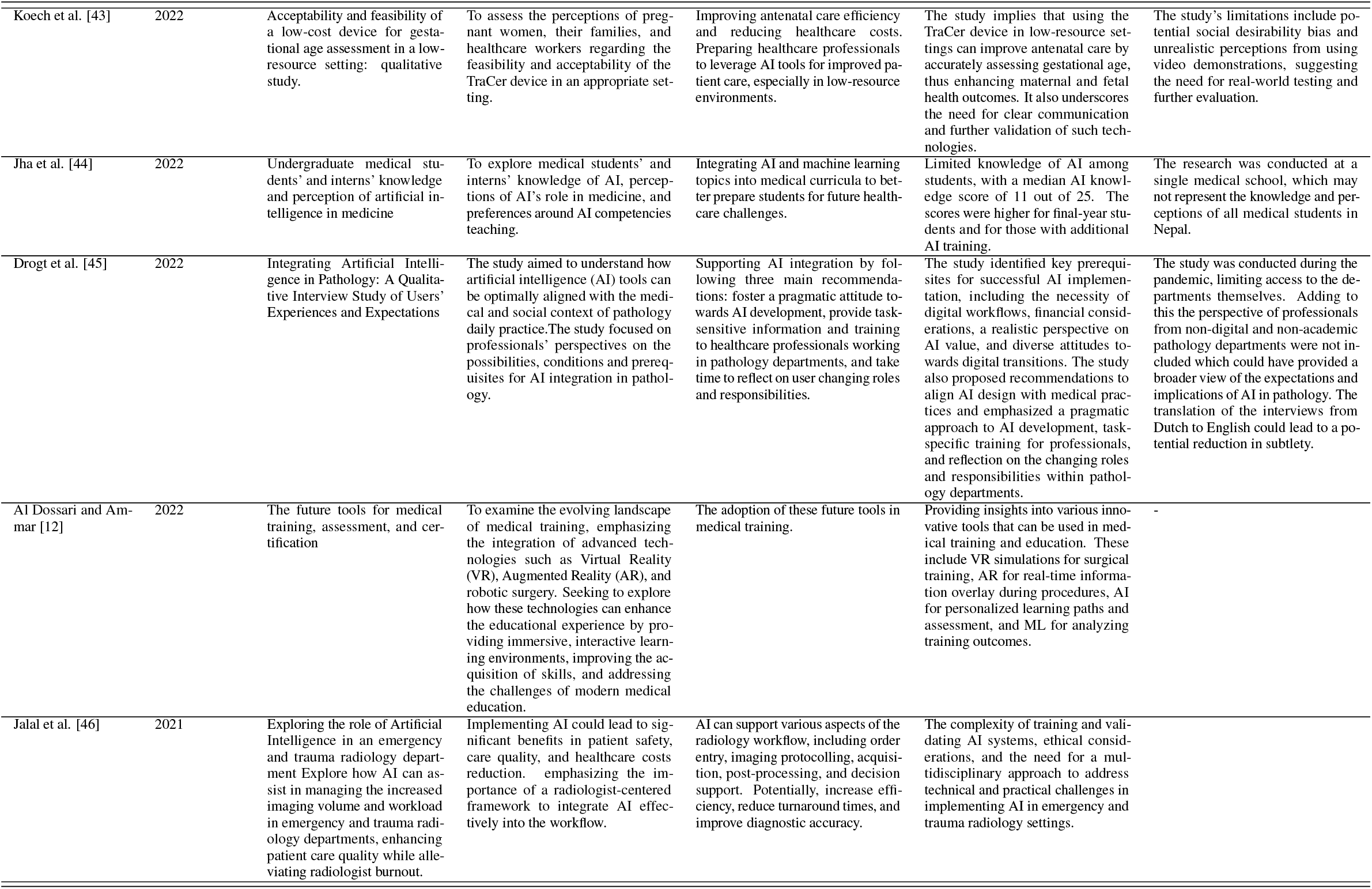
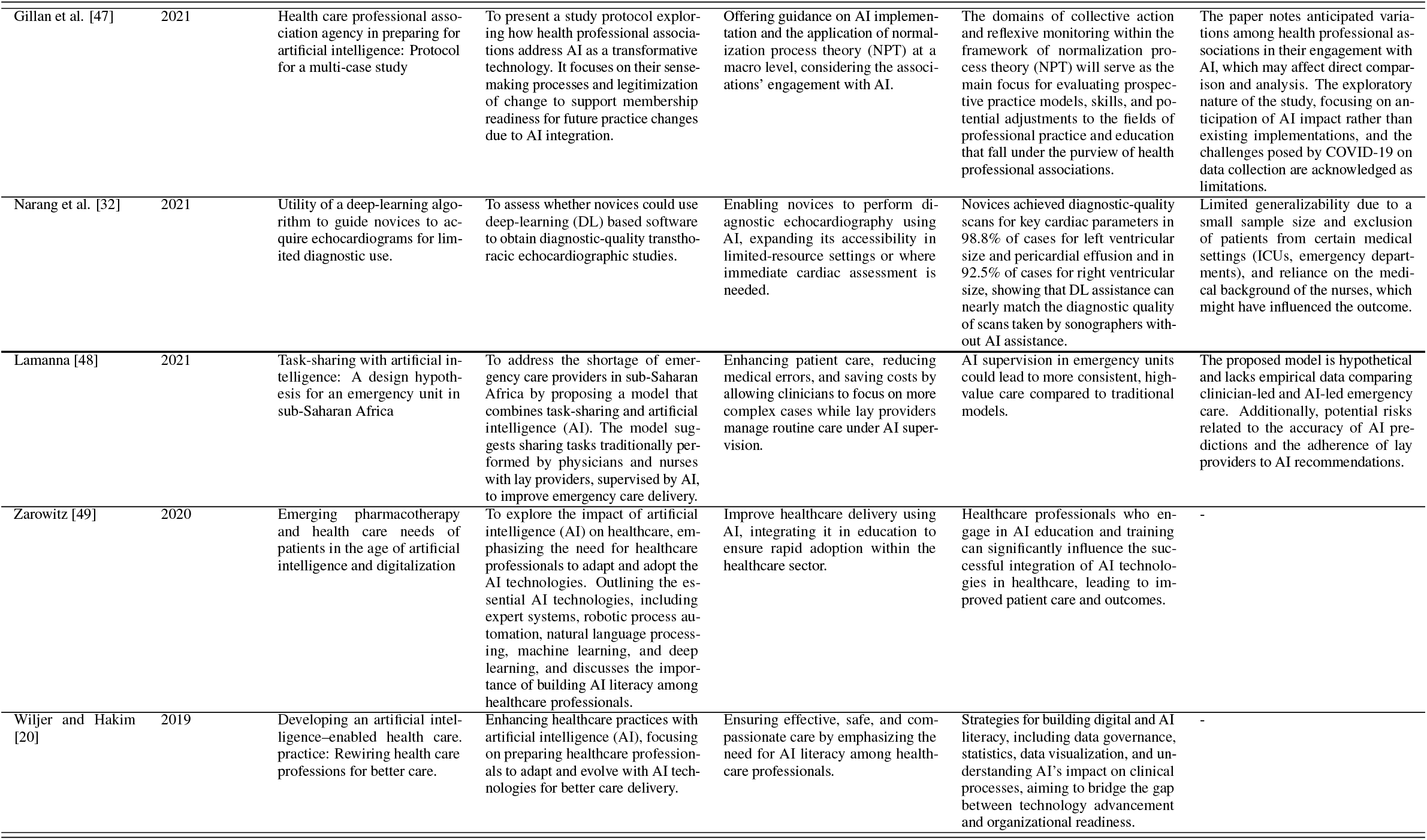
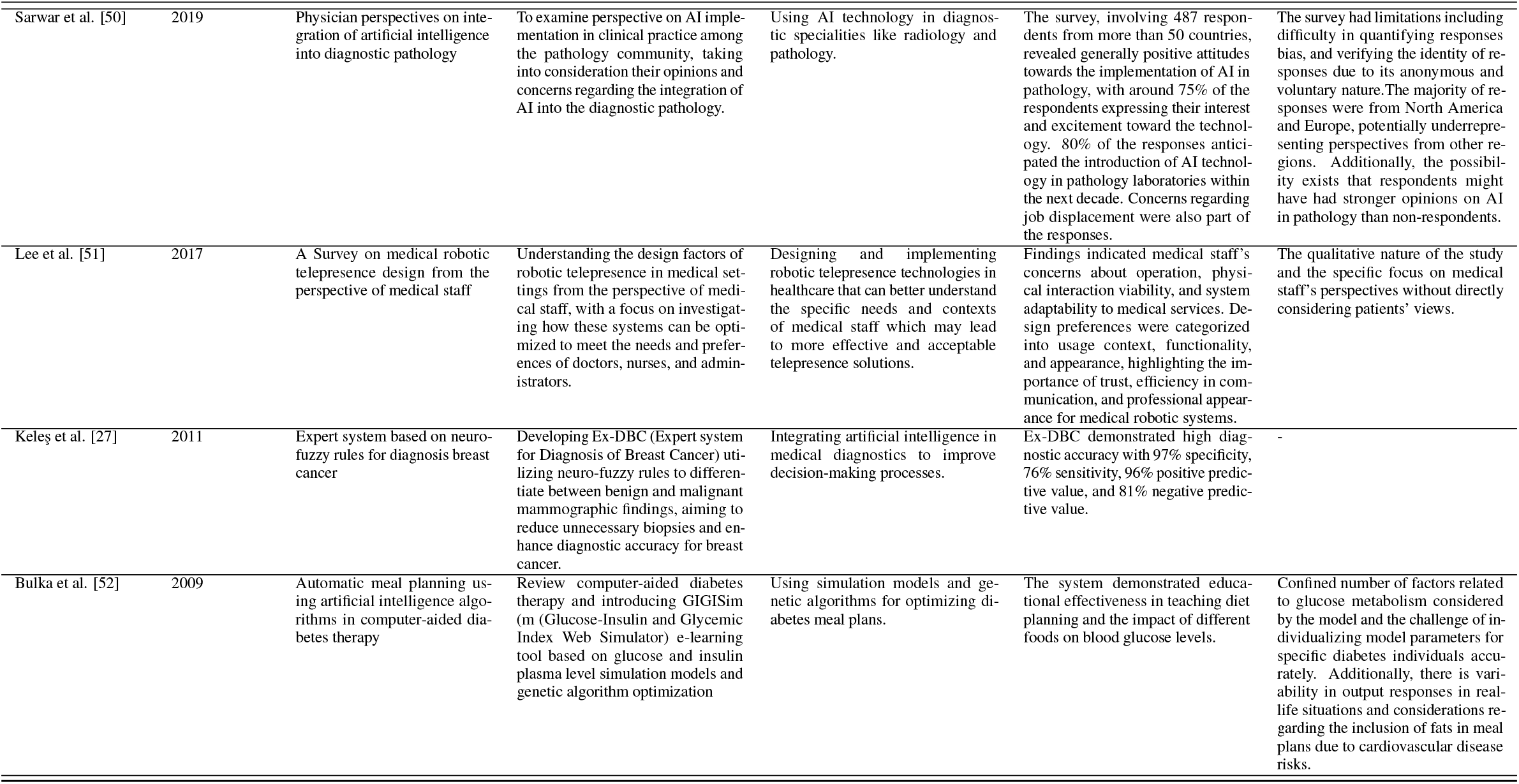
Characteristics of included studies.

### 3.1 AI as Innovative Education and Practice in Healthcare

AI is revolutionizing the training of healthcare professionals by providing innovative solutions that enhance learning and improve patient care outcomes [21]. For instance, in one study, first-year medical students reported that evaluating ChatGPT enhanced their grasp of the complexities in clinical cases [22]. In another study, integrated virtual and face-to-face simulation has been utilized to enhance clinical judgment and healthcare students’ understanding of patients’ experiences, offering a more immersive and effective training environment than traditional methods alone [14]. In one study, Truong et al. (2022)[23] have demonstrated the efficacy of AI-enhanced virtual reality simulation in preparing healthcare teams for emergency situations, such as operating room (OR) fires. Their study highlights the value of AI in creating realistic, high-stakes training scenarios that improve preparedness and confidence. Moreover, Seo et al. (2023)[24] contribute by introducing VR SBIRT training with conversational AI, providing nursing students with diverse, interactive learning scenarios [24]. This capability is crucial for preparing students to handle unexpected challenges in real-world clinical settings, ultimately leading to better preparedness and confidence in patient care. Thus, AI-based training methods are not only more adaptive and immersive but also provide a comprehensive platform for learning that traditional methods struggle to match.

Building on this foundation, AI’s influence extends to health practice, preparing professionals for future challenges with advancements in decision-making, patient monitoring, and care coordination. This progression indicates a smooth shift from education to practice, where AI continues to play a pivotal role in enhancing the capabilities of healthcare professionals [17]. Additionally, the adoption of AI tools like ChatGPT and generative models in nursing education is reshaping the field [25, 26]. These tools facilitate continuous education, provide advice, and ensure easy access to information while prioritizing ethical standards and data privacy. By integrating these tools, health education is not only about acquiring knowledge but also about understanding how to leverage technology to improve care delivery and patient outcomes.

In medical diagnostics, AI tools such as the Ex-DBC, a neuro-fuzzy rule-based expert system, are enhancing the precision of breast cancer diagnoses and reducing unnecessary biopsies [27]. AI also simplifies complex procedures for healthcare providers, exemplified by the automated assessment of left ventricular ejection fraction with ultrasound devices by oncology staff. This application streamlines cardiac evaluations, enabling efficient and accurate assessments by non-specialists and ultimately improving patient outcomes [28].

Within professional training, AI is playing a pivotal role in improving the precision and safety of clinical practices. Chen and Wang (2023) [29] have contributed to this progress by introducing an AI-assimilated Preventive Training Measure (AI-PTM) that enhances medical training for fitness nurses, focusing on the reduction of needle injury incidence and promoting preventive measures.

Given these, AI is reshaping healthcare training and practice by enhancing learning environments with advanced simulations and tools, improving both educational outcomes and patient care. These innovations can offer a seamless transition from educational settings to clinical practice, equipping professionals with essential skills for modern healthcare challenges.

### 3.2 Strategies to Foster Readiness to Use AI in Healthcare

To foster readiness to use AI among healthcare professionals, it is crucial to adopt a comprehensive approach that addresses the multifaceted dimensions of AI integration into healthcare. The necessity of integrating AI education into healthcare curricula is foundational to clarifying AI for healthcare professionals [17, 18, 20, 30]. For example, the importance of education and training in enhancing readiness for AI in healthcare settings is highlighted by Taskiran (2023) [18], who investigated the impact of an artificial intelligence course on nursing students’ readiness for medical AI. This study found that introducing AI education early in the curriculum significantly increased students’ confidence and willingness to engage with AI technologies, underscoring the necessity of embedding AI training within healthcare education frameworks. Similarly, Rony et al. (2024) [17] assessed how AI could advance nursing practice, suggesting that AI’s potential to improve decision support, patient monitoring, and care coordination necessitates a foundational understanding of AI among nursing professionals. Al-Sabawy (2023) [30] reveals that a comprehensive and accessible AI education in nursing can significantly address concerns and enhance the integration of AI into nursing practices.

The study by Hussein Mohamed et al. (2023) [31] highlights the critical role of continuous educational workshops and feedback in enhancing AI acceptance among healthcare professionals. The study found that nurses’ primary sources of information on artificial intelligence were the Internet, TV, and Facebook. After an educational program, there was a notable improvement in both their knowledge and attitudes toward AI. This emphasizes the necessity for continuous training and assistance to effectively incorporate AI into healthcare routines.

Further, hands-on experience with AI tools, builds confidence in healthcare professionals, allowing them to view AI as an augmentation of their skills. For instance, Seo et al. (2023) [24] used a virtual reality platform with conversational AI for nursing students to practice patient interactions, enhancing their clinical skills through immersive learning. Similarly, Narang et al. (2021) [32] employed a deep-learning algorithm to guide novices in performing echocardiograms, showing AI’s ability to augment medical training. These studies demonstrate the value of hands-on experience with AI tools in building healthcare professionals’ confidence and competence.

Ethical and professional integrity concerns are also paramount [33, 34, 35, 36]. Gosak et al. (2024) [25] explored nurses’ perceptions and readiness regarding AI, finding a broad recognition of AI’s role in supporting nursing work, but also a clear need for ethical guidelines to navigate the integration of AI in patient care effectively. Berşe et al. (2024) [26], highlighted that AI chatbots, like ChatGPT, have the potential to enhance the nursing educational experience by offering personalized learning opportunities to students, but stresses the need for careful consideration of ethical issues and information accuracy to protect patient care. In another study, the authors recommend prioritizing three main areas to promote responsible use of AI, including the creation of a strong ethical framework [34]. These studies underscore the necessity of addressing ethical considerations in AI deployment, highlighting that such efforts can significantly influence healthcare professionals’ acceptance of AI technologies.

Effective AI integration requires well-established infrastructure and supportive policies. Al Dossari and Ammar (2022) [12] explore innovative tools in medical training, such as virtual reality (VR) and augmented reality (AR), highlighting the need for infrastructural and policy frameworks that support the adoption of these advanced technologies. Their research suggests that immersive learning environments enabled by VR and AR can significantly enhance the medical training experience, pointing to the broader implications of supportive policies and infrastructure in facilitating the integration of AI into healthcare education.

In summary, the successful integration of AI into healthcare practices requires a strategic approach that includes education, continuous workshops, ethical considerations, and supportive policies. By addressing these key aspects, healthcare organizations can ensure readiness among healthcare professionals for the effective utilization of AI technologies, thereby enhancing patient care and operational efficiency.

## 4 Limitations

This literature review is constrained by its reliance on English-language publications and three primary databases, which might miss relevant non-English and interdisciplinary studies. The challenge of synthesizing findings from diverse methodologies limits the ability to draw generalized conclusions. Additionally, the rapid advancement of AI technology may not be fully represented, and the focus is predominantly on AI’s educational uses within healthcare, neglecting its potential impact in other crucial areas.

The reviewed studies have several limitations that impact their validity and applicability. Issues include bias from self-reported measures [14], lack of long-term follow-up [26], limited sample sizes [32], and the specific contexts of the studies which restrict generalizability [40, 45]. Additionally, many studies lack control groups or comparative analyses [14], and some fail to address ethical concerns and the implications of technological bias in AI-driven tools [26]. These limitations highlight the need for more rigorous research designs and comprehensive validation to improve the credibility and scalability of AI and simulation-based educational interventions in healthcare.

## 5 Conclusion

In summary, the integration of AI into healthcare education has been widely embraced due to its capacity to enhance diagnostic precision and simplify intricate medical procedures. Nevertheless, it is important to note the existing gaps in the literature that can guide future research endeavors.

Firstly, it is critical to develop long-term training protocols that not only evaluate the lasting impacts of AI on healthcare training but also ensure continuous updates in the curriculum to reflect rapid technological changes. Secondly, integrating the patient’s perspective into AI applications is essential for preserving a patient-centered approach in healthcare. Future investigations should focus on measuring patient satisfaction and the direct impacts of AI-trained professionals on patient outcomes. Thirdly, the advancement in explainable AI (XAI) models is also crucial. Improving the transparency of AI decision-making processes will help build trust and acceptance among healthcare providers, ensuring AI is used ethically in clinical settings. Lastly, the question of accountability in medical decisions involving AI cannot be overlooked. Establishing clear legal and ethical guidelines is imperative to navigate the complexities and maintain trust in AI-enhanced healthcare settings.

Moving forward, addressing these challenges with thorough research and strategic policy making will be pivotal in maximizing the benefits of AI for training healthcare providers and enhancing patient care outcomes.

## Data Availability

Not applicable (review paper)

## Notes

### Competing Interest Statement

The authors have declared no competing interest.

### Funding Statement

This study did not receive any funding

